# Artificial neural networks in neurorehabilitation: A scoping review

**DOI:** 10.1101/2020.02.20.20025858

**Authors:** Sanghee Moon, Pedram Ahmadnezhad, Hyun-Je Song, Jeffrey Thompson, Kristof Kipp, Abiodun E Akinwuntan, Hannes Devos

## Abstract

**BACKGROUND:** Advances in medical technology produce highly complex datasets in neurorehabilitation clinics and research laboratories. Artificial neural networks (ANNs) have been utilized to analyze big and complex datasets in various fields, but the use of ANNs in neurorehabilitation is limited. OBJECTIVE: To explore the current use of ANNs in neurorehabilitation. METHODS: PubMed, CINAHL, and Web of Science were used for literature search. Studies in the scoping review (1) utilized ANNs, (2) examined populations with neurological conditions, and (3) focused rehabilitation outcomes. The initial search identified 1,136 articles. A total of 19 articles were included. RESULTS: ANNs were used for prediction of functional outcomes and mortality (n = 11) and classification of motor symptoms and cognitive status (n = 8). Most ANN-based models outperformed regression or other machine learning models (n = 11) and showed accurate performance (n = 6; no comparison with other models) in predicting clinical outcomes and accurately classifying different neurological impairments.

**CONCLUSIONS:** This scoping review provides encouraging evidence to use ANNs for clinical decision-making of complex datasets in neurorehabilitation. However, more research is needed to establish the clinical utility of ANNs in diagnosing, monitoring, and rehabilitation of individuals with neurological conditions.

## 1. Introduction

Emerging new technologies are reshaping healthcare systems. Artificial intelligence (AI) is positioned at the front line of this transformation with the potential to provide better healthcare including accurate prediction of clinical outcomes and precise classification of diseases and symptoms (Lisboa, 2002). An artificial neural network (ANN) is a common machine learning method in AI technology. ANNs have rapidly adopted in various fields in healthcare and clinical decision-making (Shahid, Rappon, & Berta, 2019). For instance, radiologists and oncologists use ANNs in thoracic imaging (e.g., identification of pulmonary nodules and automatic categorization of benign or malignant tumor) (Hosny, Parmar, Quackenbush, Schwartz, & Aerts, 2018), brain imaging (e.g., brain tumor diagnostic prediction, imaging segmentation, benign, malignant, primary, or metastatic tumor categorization) (Orringer et al., 2017), and mammography (e.g., breast cancer detection) (Rodriguez-Ruiz et al., 2019). Cardiologists utilize ANN-based models for automatic cardiac imaging classification and electrocardiogram classification and monitoring system (Abdolmanafi, Duong, Dahdah, & Cheriet, 2017; Kiranyaz, Ince, & Gabbouj, 2016).

The ANN is inspired by biological neural processes in the central nervous system (McCulloch & Pitts, 1943; Mohri, Rostamizadeh, & Talwalkar, 2012). The primary function of neurons in the brain is to receive a signal and to deliver it to the next neuron. Similarly, ANNs also consist of simple processing elements, which are known as artificial neurons or ‘nodes’. A group of nodes 54that arrange in a parallel structure is called a ‘layer’. Figure 1 shows the three types of layers (input, hidden, output) of a simple ANN. In this ANN, the input layer receives information, which is propagated through one adjacent hidden layer to the output layer. In our brain, connections between neurons are reinforced as we repeatedly try to classify different objects or predict a consequence based on current information. Similarly, ANNs are designed to learn from data and optimize a network through reinforcing weighted connections that can classify objects or predict outcomes. The learning potential of an ANN is a major strength compared to other conventional models as the ANN can train the model to find the best subset of parameters. Depending on the complexity and types of the questions to be addressed, the number of nodes in each layer and/or the number of hidden layers can be modified. Furthermore, ANNs do not require assumption of normal distribution of variables in the dataset. As such, ANNs can compute complex and non-linear interactions between data that are often challenging to compute using conventional linear statistical analyses (Lancashire, Lemetre, & Ball, 2009). Further details about the mathematical and statistical background of ANNs can be found in the supplementary section (Supplementary Appendix S1).

**Figure 1.**
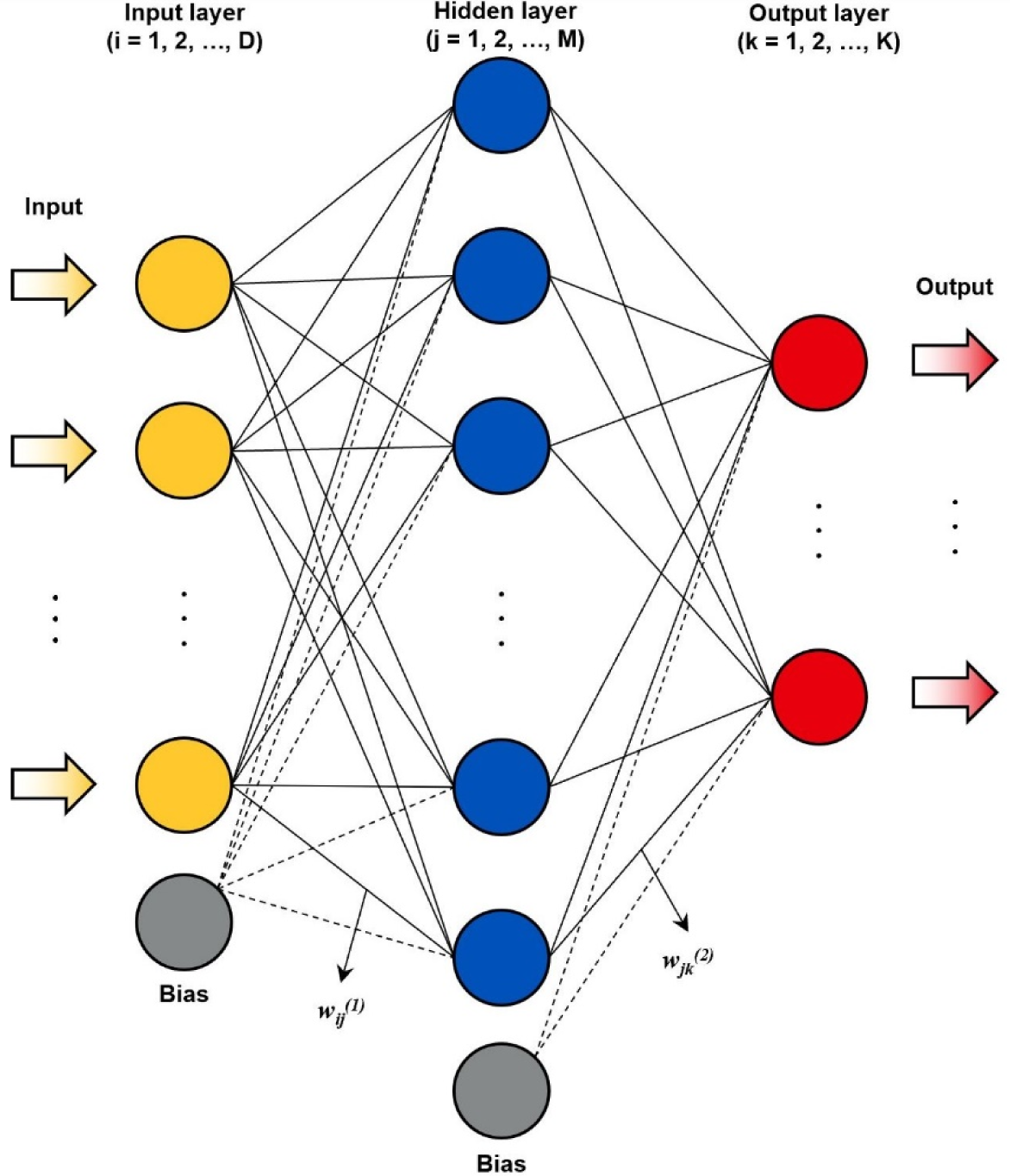
Schematic example of an ANN structure with a single hidden layer (*w*_*ij*_^*(1)*^ = weighted connection between *i*-th node in the input layer and *j*-th node in the hidden layer and *w*_*ij*_^*(2)*^ = weighted connection between *j*-th node in the hidden layer and *k*-th node in the output layer; *i* = 1, 2, …, *D*; j = 1, 2, …, M; k = 1, 2, …, K)

In neurorehabilitation, clinicians and researchers are particularly interested in predicting functional outcomes accurately and classifying subtle differences in symptoms and status correctly in order to provide better healthcare service to patients. Hence, technologically advanced measures, such as wearable gait sensors (e.g., accelerometers, gyroscopes, and magnetometers), physical activity monitors (e.g., Actigraph and StepWatch), cardiovascular health monitors (e.g., wearable electrocardiogram monitor), have seen implementation both in research and in the clinic to improve the quality of patient care (Lukowicz, Kirstein, & Tröster, 2004). These technological tools make massive volumes of quantitative data clinically available to determine health and function for patients in need of rehabilitation and community re-integration. However, these datasets are often under-utilized due to the complexity of the data and under-estimation of the potential value of data by clinicians (Ohno-Machado & Rowland, 1999).

Therefore, the purpose of this scoping review is to map the relevant literature pertaining to the use of ANNs in neurorehabilitation, with a special focus on its application for prediction of clinical outcomes and classification of clinical symptoms and status. A scoping review aims to offer a broad overview of a topic of interest, identify the literature and the feasibility of a full systematic review, include a broad range of study designs and methodologies, and identify research gaps (Arksey & O’Malley, 2005; Levac, Colquhoun, & O’Brien, 2010).

## 2. Methods

All methodological frameworks for this scoping review were primarily based on the scoping review guidelines proposed by Arksey and O’Malley (Arksey & O’Malley, 2005) including (1) identifying the research question, (2) identifying relevant studies, (3) selecting studies, (4) charting the data, and (5) collating, summarizing, and reporting the results. The Preferred Reporting Items for Systematic Reviews and Meta-Analyses (PRISMA) Extension for Scoping Reviews were utilized to improve the quality of the reporting process (Tricco et al., 2018).

### 2.1 Research question

“What is the current use of ANNs in neurorehabilitation?”

### 2.2 Identifying relevant studies

Relevant studies were identified from three databases: PubMed, CINAHL, and Web of Science. The literature search included articles from inception of each database up to September 2019. For PubMed and CINAHL, we utilized Medical Subject Headings (MeSH) terms to facilitate the literature search, but MeSH terms were not available in Web of Science. Search keyword strings were created around three main concepts to answer our research question (Table 1). The first concept was ‘ANN’. The second concept was ‘neurological conditions’, with a specific focus on the most prevalent neurological conditions (stroke, Alzheimer disease (AD), Parkinson disease (PD), spinal cord injury, and traumatic brain injury) (Feigin et al., 2017; Gooch, Pracht, & Borenstein, 2017; Pringsheim, Fiest, & Jette, 2014). The third concept was ‘rehabilitation’. Rehabilitation was defined as medical practices that involve a broad area of medical specialties and integrate various aspects of injuries and diseases that impact human functioning and health (Stucki, Cieza, & Melvin, 2007). Thus, we developed a comprehensive set of keywords revolving around rehabilitation, functioning, activities of daily living, and quality of life. We also manually searched the gray literature and the reference lists of the included studies to find additional articles. Figure 2 shows the PRISMA flowchart of the article selection process.

**Table 1.** Concepts and keywords used for literature search

| Concept | Keywords |
| --- | --- |
| Artificial neural network | Artificial neural network; *Neural Networks (Computer) [Mesh]* |
| Neurological conditions | Parkinson*; *Parkinson Disease [Mesh]*<br>stroke; *Stroke [Mesh]*<br>Alzheimer*; *Alzheimer Disease [Mesh]*<br>dementia; *Dementia [Mesh]*<br>epilepsy; *Epilepsy [Mesh]*<br>multiple sclerosis; *Multiple Sclerosis [Mesh]*<br>traumatic brain injury; *Brain Injuries, Traumatic [Mesh]*<br>Huntington*; *Huntington Disease [Mesh]*<br>amyotrophic lateral sclerosis; *Amyotrophic Lateral Sclerosis [Mesh]*<br>spinal cord injury; *Spinal Cord Injuries [Mesh]*<br>brain tumor; *Brain Neoplasms [Mesh]*<br>brain cancer; *Brain Neoplasms [Mesh]* |
| Rehabilitation | quality of life; *Quality of Life [Mesh]*<br>activities of daily living; *Activities of Daily Living [Mesh]*<br>pain; *Pain [Mesh]*<br>function<br>physical therapy; *Physical Therapy Modalities [Mesh]*<br>rehabilitation; *Rehabilitation [Mesh]*<br>assessment; *Physical Examination [Mesh]*<br>evaluation<br>treatment<br>gait; *Gait [Mesh]*<br>balance; *Postural Balance [Mesh]*<br>occupational therapy; *Occupational Therapy [Mesh]*<br>speech language pathology; *Speech-Language Pathology [Mesh]*<br>cognition; *Cognition [Mesh]*<br>memory; *Memory [Mesh]*<br>problem solving; *Problem Solving [Mesh]* |
| * was used as the truncation operator in the search. |  |
| MeSH terms (in italics) were used in addition to keywords for the PubMed and CINAHL search. |  |

**Figure 2.**
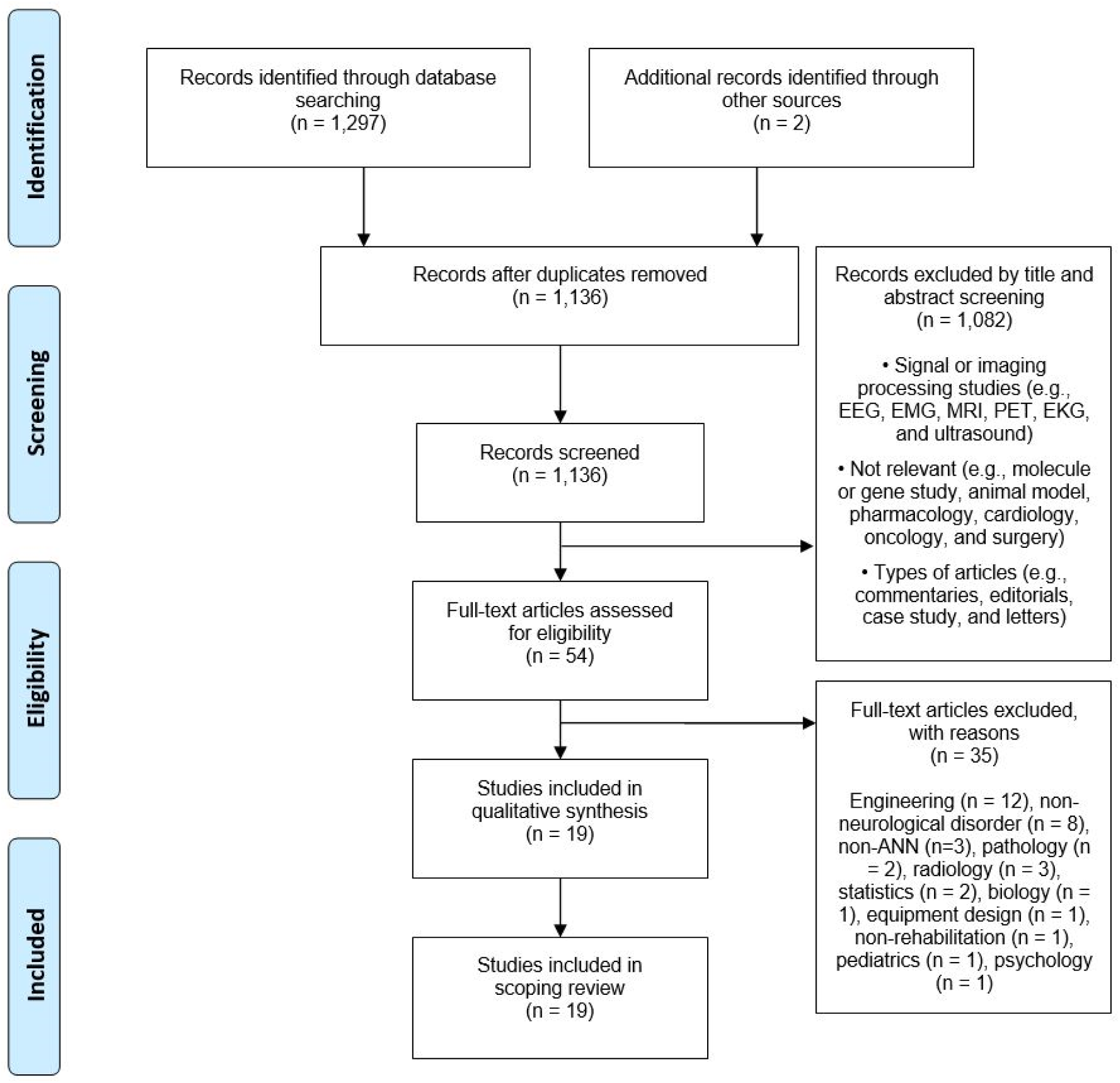
Search flow for scoping review following the PRISMA guidelines.

### 2.3 Study selection

Studies were included if they (a) utilized artificial neural networks, (b) investigated populations with neurological conditions, and (c) involved any aspect of rehabilitation as suggested in Stucki and colleagues (Refer to Section 3.2) (Stucki et al., 2007). The types of studies considered for inclusion were randomized controlled trials, retrospective database studies, cohort studies, cross-sectional studies, case reports and series, and case-control studies. Studies were excluded if they were letters, opinions, commentaries, editorials, abstracts, animal studies, reviews, or protocols. Literature written in non-English was excluded. Studies were also excluded if they did not include a functional measure or rehabilitation outcome, such as symptom severity scores, cognition assessment results, mortality rates, or gait and balance characteristics. Finally, studies that employed deep learning neural networks (e.g., convolutional neural networks and recurrent neural networks) for signal or imaging processing (e.g., electroencephalogram and functional magnetic resonance imaging) were also excluded, because simple ANNs are considered sufficient for classification and predictive modelling of most datasets (Dreiseitl & Ohno-Machado, 2002).

### 2.4 Charting the data

An Excel spreadsheet and Rayyan, a free systematic review online application, were utilized to chart the extracted data (Ouzzani, Hammady, Fedorowicz, & Elmagarmid, 2016). Titles and abstracts were reviewed by one reviewer (S.M.) and selected full texts were independently reviewed by two reviewers (S.M. and P.A.). After the independent review, there were three disagreements between two reviewers (S.M. and P.A.), which were discussed until consensus was achieved.

## 3. Results

### 3.1 Characteristics of included studies

A total of 19 studies that utilized ANNs to examine various aspects of neurological rehabilitation were used for the qualitative synthesis and included for the scoping review. Among them, 11 studies were ‘prediction’ studies, while 8 studies were ‘classification’ studies using ANNs. Most studies had a retrospective design (n = 12), followed by cross-sectional design (n = 6) and cohort design (n = 1). Descriptions of the included studies are shown in Table 2.

**Table 2.** Purposes of ANN application, patient population, and outcomes of the included studies

| **Study** | **ANN application** | **Patient population** | **Outcome of interest** |
| --- | --- | --- | --- |
| Salgueiro et al. (2013) | Prediction | Fibromyalgia | Treatment result prediction |
| Segal et al. (2006) | Prediction | TBI | Long-term outcome prediction |
| Rughani et al. (2010) | Prediction | TBI | Prediction of in-hospital mortality |
| Shi et al. (2013) | Prediction | TBI | Prediction of in-hospital mortality |
| Lu et al. (2015) | Prediction | TBI | Long-term outcome and mortality prediction |
| Pourahmad et al. (2016) | Prediction | TBI | Prognostic prediction of functional outcomes (disability) |
| Belliveau et al. (2016) | Prediction | SCI | Long-term functional outcome prediction |
| Sonoda et al. (1997) | Prediction | Stroke | Functional recovery prediction |
| Oczkowski et al. (1997) | Prediction | Stroke | Functional outcome prediction |
| Ottenbacher et al. (2001) | Prediction | Stroke | Rehospitalization prediction |
| Cheng et al. (2014) | Prediction | Stroke | Prognostic prediction of stroke recovery |
| Keijsers et al. (2000) | Classification | PD | Detection and assessment of the severity of levodopa-induced dyskinesia |
| Keijsers et al. (2003) | Classification | PD | Automatic assessment of levodopa-induced dyskinesia |
| Muniz et al. (2009) | Classification | PD (DBS) | Assessment of the effects of DBS via gait analysis |
| Manap et al. (2011) | Classification | PD | Identification of gait parameters for gait abnormality |
| Kaczmarczyk et al. (2009) | Classification | Stroke | Gait pattern classification via gait analysis |
| Scheffer et al. (2012) | Classification | Stroke | Gait pattern classification using inertial motion capture |
| Lins et al. (2017) | Classification | MCI, dementia | Identification of prognostic parameters for cognitive impairment |
| Quintana et al. (2012) | Classification | MCI, AD | Assessment of cognitive impairment using clinical neuropsychology |
| AD = Alzheimer disease, DBS = deep brain stimulation, MCI = mild cognitive impairment, PD = Parkinson disease, SCI = spinal cord injury, TBI = traumatic brain injury |  |  |  |

### 3.2 Artificial neural networks in neurorehabilitation research

In neurorehabilitation, precise prognosis and prediction of recovery and an accurate classification of the severity of diseases and injuries can enhance the quality of patient care. The studies included in this scoping review focused on prediction of long-term outcomes (Belliveau et al., 2016; Lu et al., 2015; Salgueiro et al., 2013; Segal et al., 2006), functional recovery (Cheng, Lin, & Chiu, 2014; Oczkowski & Barreca, 1997; Pourahmad, Hafizi-Rastani, Khalili, & Paydar, 2016; Sonoda, Chino, Domen, & Saitoh, 1997), mortality (Lu et al., 2015; Rughani et al., 2010; Shi, Hwang, Lee, & Lin, 2013), rehospitalization (Ottenbacher et al., 2001), and classification of functional outcomes such as movement and gait characteristics (Kaczmarczyk, Wit, Krawczyk, & Zaborski, 2009; Keijsers, Horstink, & Gielen, 2003; Keijsers, Horstink, van Hilten, Hoff, & Gielen, 2000; Manap, Tahir, & Yassin, 2011; Muniz et al., 2009; Scheffer & Cloete, 2012) and cognitive impairments (Lins et al., 2017; Quintana et al., 2012).

#### 3.2.1 Artificial neural networks to predict outcomes in neurorehabilitation

Among the 11 studies that used ANNs to predict neurorehabilitation outcomes, five studies investigated prognosis and mortality after traumatic brain injury (TBI) (Lu et al., 2015; Pourahmad et al., 2016; Rughani et al., 2010; Segal et al., 2006; Shi et al., 2013) and one study investigated prognosis and mortality after traumatic spinal cord injury (SCI) (Belliveau et al., 2016). Four studies examined functional recovery and rehospitalization after stroke (Cheng et al., 2014; Oczkowski & Barreca, 1997; Ottenbacher et al., 2001; Sonoda et al., 1997). One study used ANN to predict treatment response in people with fibromyalgia (Salgueiro et al., 2013).

Rughani and colleagues (Rughani et al., 2010) compared the accuracy of an ANN model with logistic regression in 100 patients with TBI. The ANN model predicted mortality based on clinical outcomes. The performance of the ANN model showed significantly greater accuracy (88%) in predicting survival following TBI than the logistic regression model (79%, p < 0.001) and a clinician’s judgment of survival (72%, p < 0.0001). A similar study reported that the ANN model outperformed a logistic regression model in predicting mortality with more than 95% accuracy compared to 82% in the logistic regression model (p < 0.001) (Shi et al., 2013). Lu and colleagues (Lu et al., 2015) compared the ability of multiple models including ANN, decision tree, naïve Bayes, and logistic regression to predict 6-month functional outcome and mortality in 115 people after TBI from clinical data. The ANN was the best model to predict the 6-month functional outcomes between favorable (Glasgow Outcome Scale (GOS) 1 to 3) and unfavorable (GOS 4 to 5) outcomes (area under the receiver operating characteristic curve (AUC) 96% for ANN, 94% for naïve Bayes, 92% for decision tree, and 90% for logistic regression), but it was not the best model to predict 6-month mortality. A naïve Bayes model (AUC 91%) outperformed other models (AUC 81% for ANN, 78% for decision tree, and 87% for logistic regression) in predicting 6-month mortality after TBI.

To predict functional outcomes after TBI, two studies utilized ANN models. An ANN-hybrid model with a decision tree predicted 6-month post functional outcomes (extended Glasgow Outcome Scale) in 410 people after TBI with an 86% accuracy, whereas the decision tree model yielded 82% (Pourahmad et al., 2016). Contrarily, Segal and colleagues (Segal et al., 2006) reported the ANN model not to show greater accuracy (r^2^ = 0.40 between predicted and observed outcomes) in predicting 1-year post-TBI outcomes such as Functional Independence Measure (FIM; total score of motor and cognitive subscales), Disability Rating Scale, and Community Integration Questionnaire, compared with multiple regression models (r^2^ = 0.44) and classification and regression trees (r^2^ = 0.29).

One study utilized ANNs to predict outcomes in the SCI population. Belliveau and colleagues (Belliveau et al., 2016) analyzed the data of 3,142 people with traumatic SCI who were registered in the national spinal cord injury model systems database. An ANN model was used to predict the level of independence one year after discharge from hospital. The ANN model predicted the ambulation status (e.g., 150ft walk, 1 street block walk, and 1 flight of stairs) with a high accuracy (85.5 to 87.7%) and the non-ambulation outcomes (e.g., bed-chair transfers, bowel and bladder management, eating, and toileting) with a high accuracy (75.9% to 83.4%). However, their ANN model performed similarly compared to the logistic regression model (86.0 to 87.9% for ambulation status and 75.6 to 85.9% for non-ambulation status).

Another study investigated 72 people with fibromyalgia to evaluate the accuracy of ANNs to predict treatment response (responder vs. non-responder) after a 4-week interdisciplinary pain program at discharge and 6-month follow-up (Salgueiro et al., 2013). Functional outcomes such as the Stanford Health Assessment Questionnaire and the McGill Pain Questionnaire were used as input data. Compared to logistic regression models, ANN models showed superior prognostic accuracy in predicting treatment response at discharge (91.7% for ANN and 86.1% for logistic regression) and at 6-month follow-up (91.7% for ANN and 61.1% for logistic regression).

Three studies used ANN models to predict functional outcomes and one study to predict rehospitalization after stroke. Oczkowski and Barreca (Oczkowski & Barreca, 1997) tested an ANN model to predict functional recovery in 147 stroke survivors. The ANN model predicted discharge FIM score (total score of motor and cognitive subscales) with 88% accuracy, but this study did not have any comparison model. Sonoda and colleagues (Sonoda et al., 1997) used an ANN model to predict functional outcomes in 70 people after stroke using the severity of impairment. In this study, the predicted FIM scores by the ANN model and observed FIM scores were significantly correlated (*r* = 0.74; p < 0.001), but no comparison model was reported.

Cheng and colleagues (Cheng et al., 2014) developed an ANN model to predict prognosis 3 months after ischemic stroke based on the data of 82 ischemic stroke survivors. Their model showed greater accuracy (95%) in predicting prognostic outcomes compared to the logistic regression model (68%). Lastly, a large medical record database was studied by Ottenbacher and colleagues (Ottenbacher et al., 2001), which contained nearly 10,000 patient records collected from 167 hospitals across 40 states in the United States. The study compared ANN with a logistic regression model in predicting hospital readmission after stroke. This study reported no statistically significant difference or practical advantage in performance characteristics of the two models, though the ANN model (AUC 74%) showed slightly accurate performance in rehospitalization prediction than the logistic regression model (AUC 68%).

#### 3.2.2 Artificial neural networks to classify outcomes in neurorehabilitation

Eight studies utilizing ANNs primarily for classification were included in this scoping review. Among these, six studies used ANNs for classification of gait pattern or movement abnormality (Kaczmarczyk et al., 2009; Keijsers et al., 2003; Keijsers et al., 2000; Manap et al., 2011; Muniz et al., 2009; Scheffer & Cloete, 2012), while two studies used ANNs for classification of cognitive status (Lins et al., 2017; Quintana et al., 2012).

Four studies utilized ANNs for classification of motor symptoms such as dyskinesia or abnormal gait among people with PD (Keijsers et al., 2003; Keijsers et al., 2000; Manap et al., 2011; Muniz et al., 2009). Keijsers and colleagues (Keijsers et al., 2000) collected wearable accelerometer data from 16 people with PD. The ANN models differentiated voluntary movement from levodopa-induced dyskinesia, and correctly classified the severity of dyskinesia that corresponded to the modified Abnormal Involuntary Movement Scale (mAIMS; 0-4 scale; 0 = no dyskinesia and 4 = extreme dyskinesia). In this study, their ANN models showed correlations (*r*) ranging from 0.66 to 0.84 between predicted and observed mAIMS scores. The magnitude of correlations derived from the ANN was higher compared to logistic regression (*r* between -0.01 and 0.77). A follow-up study by the same group developed an automatic assessment system for levodopa-induced dyskinesia, in which the ANN correctly classified the severity of levodopa-induced dyskinesia (93.7% for arms, 99.7% for trunk, and 97.0% for legs) and distinguished dyskinesia from voluntary movements in a home-like setting (80% accuracy on average). In this follow-up study, no comparison model was reported (Keijsers et al., 2003).

Finally, ANN classifiers were used to classify gait patterns among people with PD. Their ANN model identified abnormal gait in people with PD from healthy controls with an accuracy of up to 96%, but no comparison model was utilized (Manap et al., 2011). Furthermore, another study examined the effects of deep brain stimulation on gait mechanics and reported that the ANN-based model classified people with PD and healthy controls based on their ground reaction force patterns (AUC 99.5%). No comparison model was used (Muniz et al., 2009).

Two studies using ANNs investigated gait performance in stroke survivors (Kaczmarczyk et al., 2009; Scheffer & Cloete, 2012). Scheffer and Cloete (Scheffer & Cloete, 2012) used an inertial motion capture system to measure gait mechanics during a 10-m walk at normal walking pace and reported that the ANN model accurately differentiated between stroke survivors and healthy controls based on hip, knee, and ankle gait parameters with an accuracy of 99.4%. Kaczmarczyk and colleagues (Kaczmarczyk et al., 2009) reported that the ANN model classified three types of gait patterns, based on position of the foot (forefoot, flatfoot, and heel) during first contact on the ground, among people after stroke. Their ANN model used knee and hip joint angle changes during gait cycle as inputs and classified three gait patterns with high accuracies (94% to 100%), compared with traditional analytic models such as cluster analysis and discriminant function analysis (39% to 67%).

ANNs were also used for classification of cognitive impairment such as mild cognitive impairment (MCI) and dementia. Quintana et al (Quintana et al., 2012) developed an ANN model that classifies MCI and AD based on neuropsychological profiles. In their study, the ANN model (98 to 100%) outperformed a linear discriminant analysis (80 to 96%) in classification of cognitive status. On the other hand, Lins and colleagues (Lins et al., 2017) developed regression models using different machine learning methods such as ANNs and random forest in order to classify cognitive status among normal, MCI, and dementia patients using clinical and neuropsychological outcomes. The study reported that random forest-based model showed more accurate classification (96.3 to 99.5%) compared with the ANN-based model (56.2 to 79.4%).

## 4. Discussion

In neurorehabilitation, accurate prediction of clinical outcomes and classification of different symptoms and diagnoses can inform the clinical decision-making process to provide effective and adequate treatment to patients. This scoping review evaluated the current literature on the use of ANN in neurorehabilitation. Although this review focused on various neurological conditions, study designs, study outcomes, and statistical models, our findings demonstrated that ANNs typically outperform traditional statistical methods to predict or classify clinical outcomes in neurological conditions.

One possible reason for the superior accuracy of ANNs compared to traditional regression models is the highly complex and linear datasets used in neurorehabilitation research. Accuracy of multiple linear regression and linear classification often decreases when analyzing non-linear data (Landi, Piaggi, Laurino, & Menicucci, 2010; Tu, 1996). In addition, data collected from rehabilitation clinics often have missing or corrupted variables due to human errors or unknown technical glitches. Unlike traditional methods, ANNs are robust and remain functional despite missing, incomplete, or noisy data using minimal modifications of ANNs (Sharpe & Solly, 1995; Smieja, Struski, Tabor, Zieliński, & Spurek, 2018).

The results of our scoping review suggest that the use of ANN should be considered more in neurorehabilitation research. Incorporating ANNs into clinical environments may increase the efficiency of rehabilitation practices and allow clinicians to predict disease progression and functional recovery more quickly and accurately. A non-exhaustive list of future implications includes: (1) classification of gait patterns that allow to differentiate between several neurological conditions. Individuals with neurological conditions often have impaired gait and balance, which increases the risk of falls (Homann et al., 2013). Classification of the type of gait abnormalities may assist in developing a tailor-made fall prevention and treatment program.

ANNs can be used to analyze data gathered from wearable devices (e.g., wearable inertial motion sensors) for gait assessment, often highly complex and inter-related; (2) classification of neurodegenerative changes before the onset of dementia, allowing the development of early treatment plans to slow down the disease progression. Timely diagnosis and early intervention is crucial for dementia (Robinson, Tang, & Taylor, 2015). ANN-based models may help accurately predict patients’ disease progression and classify patients at different stages. (3) Prediction of the likelihood of favorable functional outcome after brain injuries. ANNs can be utilized as an additional tool to assist prognostic decision-making process. Proper estimation of functional or recovery outcome helps to offer the most appropriate rehabilitation by clinicians. ANNs can also provide accurate estimation regarding medical cost based on the estimation of functional outcomes.

However, despite many potential benefits, ANNs still have several limitations. The most criticized limitation of ANNs is the black box problem. In ANNs, all computational processes in the hidden layer are difficult to interpret by the human users. However, several methods including neural interpretation diagram, Garson’s algorithm, and sensitivity analysis help to understand the black box problem (Olden & Jackson, 2002). A recent study developed an ANN that can show its reasoning process to the users (Mascharka, Tran, Soklaski, & Majumdar, 2018). These advancements may eventually help overcome the black box problem in the near future. In addition, small datasets can be overfitted by any statistical method including ANNs. Overfitting means that the model is optimized to the respective training dataset at the expense of the generalizability of the model. This can negatively impact the performance of the model on a new dataset. By training the ANN model with more data or by setting up early stopping and validation checks, the overfitting problem can be reduced. Lastly, the architecture of ANNs often requires subjective decisions by developers, since there is no specific guideline to construct a structure of ANN (Tu, 1996). However, this problem can be remediated by several approaches such as finding improved structures in an unbiased fashion (hyperparameter optimization) (Diaz, Fokoue-Nkoutche, Nannicini, & Samulowitz, 2017) and systematically achieving convergence with different error goals, learning rates, and number of hidden layers or number of neurons in the hidden layer (da Silva, Spatti, Flauzino, Liboni, & dos Reis Alves, 2017).

Finally, this review has limitations. In this scoping review, it was inherently difficult to conduct a comprehensive synthesis and make strong recommendations on the use of ANN in neurorehabilitation due to the heterogeneity of selected articles. However, the purpose of a scoping review is to emphasize comprehensive coverage, rather than on a particular standard of evidence (McColl et al., 2009). Second, we possibly omitted other relevant studies due to the selection of search database and the exclusion of non-English articles. However, as other scoping reviews noted (Cameron, Tsoi, & Marsella, 2008; Gentles, Lokker, & McKibbon, 2010; Levac, Wishart, Missiuna, & Wright, 2009), it may be impossible to deal with all relevant studies as scoping reviews cover a broader focus and are not designed to be as exhaustive or comprehensive.

## 5. Conclusion

This scoping review discussed the basics of ANNs including the mathematical representation of ANN structure, learning algorithm, and general data processes (Supplementary Appendix S1), and discussed current and future uses of ANNs in neurorehabilitation. Although ANNs typically showed better performance compared to traditional analytic methods in analyzing complex data, they have not been used much in neurorehabilitation research. ANNs have distinct advantages, are useful in neurorehabilitation, and have great potential to aid clinicians’ decision-making, predict accurate clinical outcomes, and classify subtle symptoms and diagnoses in neurorehabilitation clinics.

## Data Availability

Not Applicable.

## 7. Acknowledgements

None.

